# Non-Medical Management of Hypertension and Its associated Factors among Hypertensive Patients Attending Public Hospitals in Hawassa City, Ethiopia

**DOI:** 10.1101/2024.03.13.24304244

**Authors:** Meheret Mekonnen, Merihun Eshetu, Feleke Tefessse, Dereje Geleta

## Abstract

**Background:** Hypertension is a major health issue, affecting the young population worldwide. A non-medical management approach can decrease the effects of high blood pressure levels in hypertensive patients. Even though NMM is one of the most effective ways to prevent & control hypertension, only little emphasis has been given to it compared with treating hypertension with medication. Therefore, this study aimed to assess the practice of non-medical management of hypertension and associated factors among hypertensive patients attending selected public hospitals in Hawassa City, Sidama, Ethiopia.

**Method:** Facility based cross-sectional study was conducted from April-May 2023. A simple random sampling technique was used to select study participants, and data was collected using an interviewer-administered questionnaire. Data was entered into Epi-Data version 4.2 and exported into SPSS version 26 for analysis. Binary logistic regression was used to identify the association between dependent variables and independent variables.

**Result:** The study revealed that 62.3 % of participants had good practice of non-medical management of hypertension (95%CI: 56.5-67.3). Additionally, 74.7 % had good knowledge, (AOR=2.054; 95% CI: 1.180-3.575, P-value=0.011) and 65% exhibited favourable attitudes towards non-medical management of hypertension (AOR =2.368; 95% CI: 1.354-4.141, p-value=0.003). Factors such as good knowledge, rural residence (AOR=1.939 95% CI, 0.943, 3.987, P-value=0.072) and having a family history of hypertension (AOR=1.764; 95% CI: 1.073-2.899, p-value=0.025) were statistically significant in association with non-medical management practice.

**Conclusion:** This study found that nearly two-thirds of participants demonstrated good practice of non-medical management of Hypertension. Good knowledge, favourable attitude towards non-medical management, rural residence and having a family history of hypertension were significantly associated with non-medical management of hypertension.

Therefore, public health interventions focusing on non-medical management of hypertension and strengthening NCD control programs are essential. Moreover, the provision of targeted Health education and behavioural change communication to patients can improve disease knowledge and improve Non-medical management practices.

## Background

Hypertension is defined as when increased blood pressure &the force of blood flowing through blood vessels are consistently too high. This occurs when systolic blood pressure is greater than or equal to 140 mmHg or diastolic blood pressure is greater than or equal to 90 mmHg. So that, hypertension is when blood pressure is reading 140/90 mmHg(1). High blood pressure, also known as hypertension, is a major contributor to the global disease burden and is responsible for 17.9 million deaths each year globally. However, it remains widely undetected, undertreated and poorly controlled. Moreover, the number of people with uncontrolled hypertension has increased to around 1.13 billion worldwide in the past three decades(2)World Health Organization recommend evidence-based CVD prevention and management with a strategy called the Best Buys’’ strategy (WHO, 2013-2020) (3) From this, the third objective of the strategy is reducing modifiable risk factors for hypertension. On the other hand, according to global Heart initiatives, there are five technical packages. Out of this, the first package is health system strengthening which includes a healthy lifestyle, evidence-based treatment protocol, access to essential medicines and technology, risk-based management team care, task sharing and a system for monitoring. (4) Generally, hypertension should be managed in an integrated fashion with a comprehensive set of non-pharmacological and pharmacologic strategies. The main part of the non-pharmacological strategy is non-medical management (1). It includes the Dietary Approaches to Stop Hypertension (DASH) diet (high in fruits, vegetables, low-fat dairy products, potassium and calcium; low in cholesterol and saturated fat), keep dietary sodium below 100 mmol (2.4 g) daily, keep daily alcohol intake below 1 oz. (30 mL; 2 drinks) in men or 0.5 oz. (15 mL) in women, aerobic exercise for at least 30 minutes daily, achieve and maintain normal body weight, and recommend that smokers stop smoking. Compliance with all these strategies, a patient can lower blood pressure significantly and perhaps eliminate their hypertension.

## Methods and Materials

### Study setting

The study was conducted in Sidama regional state, Hawassa city, Sidama regional state, from April to May 2023. Hawassa is the capital city of Sidama Regional State. The city is located on the shores of Lake Hawassa in the Great Rift Valley at a distance of 275 km South of Addis Ababa. It covers 50.24 square kilometres, which has 8 sub-cities and 32 kebeles. Based on the information obtained from the plan and development core process of Hawassa City Administration, the city has an estimated total population of 351,469 out of which 190,757 were males and 160,712 were females (CSA, 2016). In the city, there are E11 Health centers and 4 Public Hospitals (Hawassa City Administration Health Department, 2023).

### Study participants & eligibility criteria

The study population were all Hypertensive patients who were attending their follow-ups at selected public hospitals during the study period. A randomly selected and those who fulfilled the inclusion criteria were included in the study. Adult hypertensive patients aged >18 years, attending follow-up at selected public hospitals for at least 6 months and more were included in the study. Patients who were seriously ill, having difficulties hearing and speaking (communication challenges), and patients who did not volunteer were excluded.

### Sample size determination

The sample size was calculated using a single population proportion formula by assuming that 68.92% of hypertensive patients have good practice for NMM of HTN (7) with a 95% confidence interval and 5% margin of error which is 270. Then by considering 10% of the non-response rate, the final sample size of 300 was used in the study.

### Sampling procedure and techniques

All 4 public Hospitals namely, Adare General Hospital, Motite Furra Primary Hospital, Tula Primary Hospital and Hawassa University Comprehensive Specialized Hospital located in Hawassa City were included in the study. The calculated sample size was proportionally allocated to all Hospitals based on the number of hypertension patients on the following. Study subjects (adult hypertensive patients) with hypertension attending selected public hospitals for medical follow-up were recruited by using a simple random sampling method by considering patient flow as random.

### Data collection tools and procedures

Data was collected through interviewer-administered & structured questionnaires which are adapted from previous similar studies. It contains information about socio-demographic characteristics and comprehensive practices about non–medical management of hypertension & associated factors.

Non-medical management practices were assessed using items that had Yes or No responses. The common NMM practices and activities like limiting salt consumption, engaging in regular exercise, maintaining a healthy diet, quitting smoking, and limiting alcohol consumption, consumption of caffeine, khat chewing, and regularly checking BP & weight were assessed with specific questions for participants to respond from the alternatives.

To assess the extent of non-medical management practices for hypertension among hypertensive patients, we administered a set of 15 practice-related questions. After collecting responses, we aggregated the scores of each respondent based on their answers to these questions. Those who scored at or above the mean value were categorized as having “Good practice”. While individuals who scored below the mean were classified as having,” poor practice” concerning NMM for hypertension.

To assess the level of knowledge regarding the practice of NMM for hypertension among study participants we used a set of 13 knowledge-based questions. The respondents’ answers to these questions were then analyzed by calculating the mean score, and subsequently, their knowledge scores were aggregated. Based on the cumulated scores, participants who scored above or equal to the mean were classified as having a “Good knowledge level” concerning NMM of hypertension, while those who scored below the mean were classified as having a “poor knowledge level”.

To assess the level of attitude towards the practice of NMM (Non-Medical Management) for hypertension among study participants, a questionnaire containing 11 statements was utilized. Respondents were asked to indicate their level of agreement, ranging from “strongly agree” to “strongly disagree” on a five-point Likert-type scale. After collecting and analyzing the responses, participants who scored at or above the median value were categorized as having a “favourable attitude,” while those who scored below the median were classified as having an “unfavourable attitude” towards NMM of hypertension.

The data collection tools were initially developed in English and later translated into Amharic. To ensure accuracy and desired results, we did a back translation of the instruments back into English. Before the main data collection process, a pre-test was conducted one week prior, involving 5% of the sample size, to assess the understandability and applicability of the instruments. After analyzing the pre-test data, any ambiguous or unclear questions were rephrased to enhance their understandability(10).

### Data processing and analysis

Following the data collection, each questionnaire was checked for completeness, missed values and unlikely responses and cleaned up before leaving the study area. Data was entered into Epi-data and exported to SPSS for further analysis. Descriptive statistical analysis such as frequency distribution, means, and the measure of dispersion was used to describe data and analytical statistics including bivariate and multivariable logistic regression analysis. Binary logistic regression was done to examine the association between dependent and independent variables. After running binary logistic regressions, all variables with *p* < 0.25 was considered a candidate for the final model and the corresponding *p*-value of <0.05 was considered statistically significant. Model fitness was checked by the Hosmer and Lemshow significance test that were 0.34 and multicollinearity between independent variables was checked by VIF. AOR at 95% CI was considered to declare the dependent effect of independent variables on the outcome.

### Ethical Consideration

Ethical clearance was obtained from the Institutional Review Board, (***REF.No:IRB 222/15***) of Hawassa University College of Medicine and Health Sciences. A letter of support was obtained from Hawassa University College of Medicine and Health Sciences and the Hawassa City Administration Health Department. verbal informed Consent was obtained from each study participant after informing them of all the purposes, benefits, risks and the confidentiality of the information and the voluntary nature of the participation in the study.

## Result

### Socio-demographic characteristics of the respondents

A total of 300 participants were involved in the study. The mean age of participants was 55.45±11.27 SD years and most of, 55% (n=165) of participants were aged above 55 years. From the study participants, 63.6 % (n=191) of respondents were male. Of the study participants, 83.3% (n=250) were married, and 81% (n =243) were from urban areas. More than one-third, 34.3 (n=103) of the respondents were government employees, (Table-1)

**Table 1:** Socio-demographic characteristics of the study participants in Hawassa CitY, Ethiopia (n=300)

| Variables | Frequency | Present (%) |
| --- | --- | --- |
| **Sex** |  |  |
| Male | 191 | 63.6 |
| Female | 109 | 36.3 |
| **Marital status** |  |  |
| Married | 250 | 83.3 |
| Single | 33 | 11.0 |
| Divorced | 8 | 2.7 |
| Widowed | 9 | 3.0 |
| **Age** |  |  |
| <35 years | 27 | 9.0 |
| 35-44 years | 76 | 25.3 |
| 45-54 years | 32 | 10.7 |
| >=55 years | 165 | 55.0 |
| **Permanent resident** |  |  |
| Urban | 243 | 81.0 |
| Rural | 57 | 19.0 |
| **Occupation** |  |  |
| Government employee | 103 | 34.3 |
| Private/NGO employee | 24 | 8.0 |
| Merchant | 52 | 17.3 |
| Farmer | 27 | 9.0 |
| Student | 13 | 4.3 |
| Housewife | 47 | 15.7 |
| Daily labourer | 6 | 2 |
| Retired | 28 | 9.3 |
| **Religion** |  |  |
| Orthodox | 99 | 33.0 |
| Protestant | 140 | 46.7 |
| Muslim | 47 | 15.7 |
| Catholic | 13 | 4.3 |
| Others | 1 | 0.3 |
| **Educational Status** |  |  |
| Never attended school | 15 | 5.0 |
| Can read & write | 34 | 11.3 |
| Primary school (Grades 1-8) | 38 | 12.7 |
| Secondary school (Grade 9-12) | 64 | 21.3 |
| Diploma | 83 | 27.7 |
| First Degree & above | 66 | 22.0 |
| **Income per month (Ethiopian birr)** |  |  |
| <500 | 74 | 24.7 |
| 500-1000 | 3 | 1.0 |
| 1001-1500 | 4 | 1.3 |
| >1500 | 219 | 73.0 |
| **Time since diagnosis** |  |  |
| <1year | 29 | 9.7 |
| 2-5years | 158 | 52.7 |
| 6-15years | 110 | 36.7 |
| >=15years | 3 | 1.0 |

### Practices of non-medical management of hypertension of the respondents

Among the study participants, 62.3% demonstrated good practice in non-medical management of hypertension (95%CI: 56.5-67.3). Of the study participants, 31.3%(n=94)regularly checked their blood pressure, 49% (n=147) regularly checked their weight, 81.7 % (n=245) didn’t smoke cigarettes, 75.3 % (n=226) abstained from drinking alcohol, 91.7% (n=275) did not chew khat, 33.7(n=101) refrained from drinking coffee, 77.0 % (n=231) included fruits and vegetables in their diet, 57% (n=171) Avoided consuming animal fat such a butter, and fatty meat),77%(n=231) did not use extra salt in the food. Moreover, a significant portion of the respondents, 58.3% (n=175) had experienced regular physical exercise. (Table-2)

**Table 2:** Practices of non-medical management of hypertension of the study participants in Hawassa City, Ethiopia, 2023 (300)

| Variables | Frequency | Present (%) |
| --- | --- | --- |
| Check your blood pressure regularly |  |  |
| Yes (1) | 94 | 31.3 |
| No (0) | 206 | 68.7 |
| Check your weight regularly |  |  |
| Yes(1) | 147 | 49.0 |
| No(0) | 153 | 51.0 |
| Smoke cigarette, currently |  |  |
| Yes(0) | 55 | 18.3 |
| No(1) | 245 | 81.7 |
| Drink alcohol, currently |  |  |
| Yes(0) | 74 | 24.7 |
| No(1) | 226 | 75.3 |
| Chew Khat, currently |  |  |
| Yes | 25 | 8.3 |
| No | 275 | 91.7 |
| Drink coffee, currently |  |  |
| Yes | 199 | 66.3 |
| No | 101 | 33.7 |
| Eat fruits and vegetables |  |  |
| Yes(1) | 231 | 77.0 |
| No(0) | 69 | 23.0 |
| Eat animal fat (butter, fatty meat) |  |  |
| Yes(0) | 129 | 43.0 |
| No(1) | 171 | 57.0 |
| Use extra salt |  |  |
| No | 69 | 23.0 |
| Yes | 231 | 77.0 |
| Have experience of regular physical exercise |  |  |
| Yes(1) | 175 | 58.3 |
| No(0) | 125 | 41.7 |
| Overall practice (Computed) |  |  |
| Good | 187 | 62.3 |
| Poor | 113 | 37.7 |

### Attitude towards practices of non-medical management of hypertension of the respondents

Of the study participants, 65% (195) had favorable attitudes with 95% CI (59.4-69.8). Specific to the attitudes of the study participants, 217(72.3%) strongly agreed that follow-up education on the non-medical management of hypertension is an essential component of HTN management, 140(46.7%) strongly agreed that regular checks of BP areare an important part of HTN management, 207(69.0%) were strongly agree on controlling salt intake was important for controlling blood pressure, 140(46.7%) were strongly agree on maintaining normal body weight was important for controlling blood pressure, 143(47.7%) were strongly agree on controlling diet was important for controlling blood pressure,128(42.7%) were agree on drinking caffeine affects controlling blood pressure, 138(46%) were agreed on it’s good to have whole fruits and vegetables to lower blood pressure for HTN patients. On the other hand, 96(32%) strongly disagreed that smoking cigarettes does not affect blood pressure, while 116(38.7) strongly agreed that Khat chewing has an effect on the management of hypertension, and 154(51.3%) strongly agreed thatthat regular physical exercise is important in the management of hypertension (Table-3)

**Table 3:**
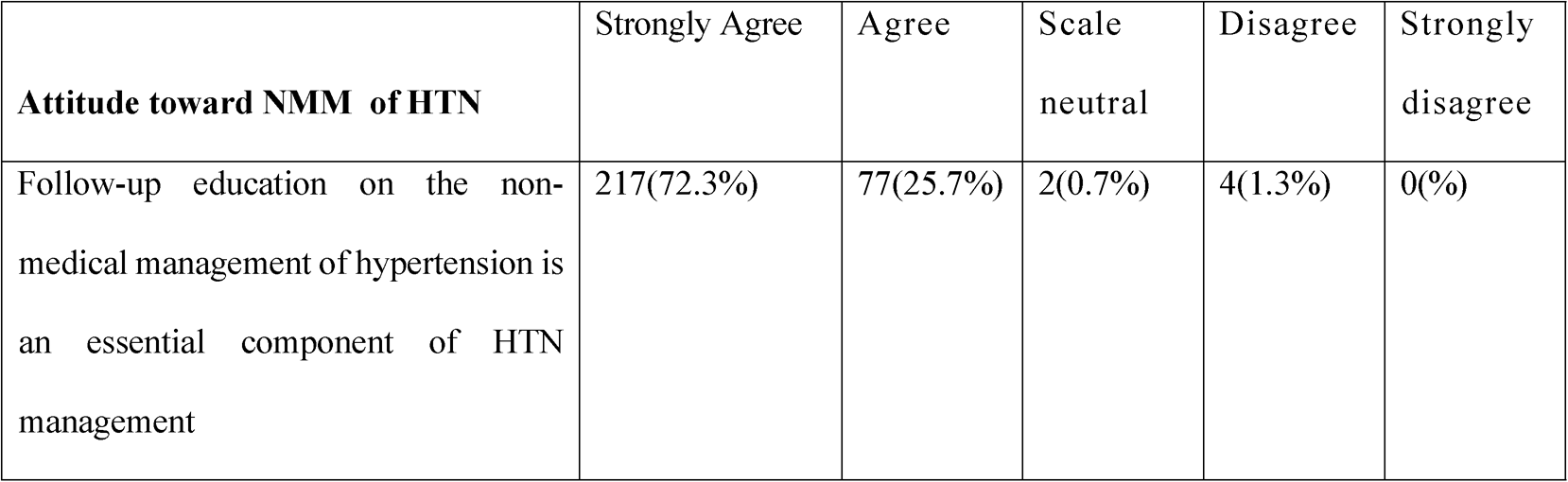
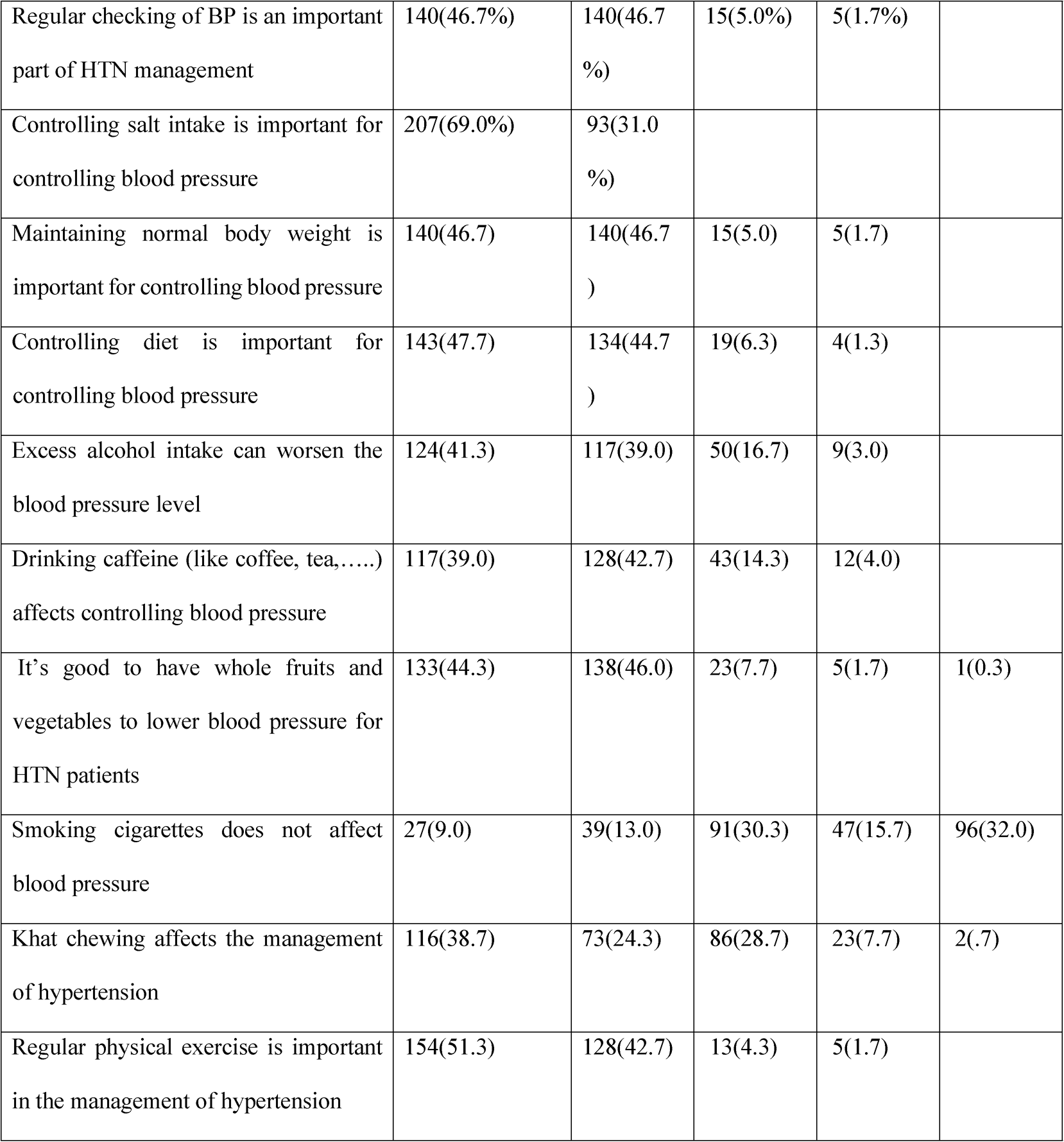
Attitude towards practices of non-medical management of hypertension of the study participants in Hawassa City, Ethiopia, 2023 (n=300)

**Table 4:** Knowledge level of respondents toward non-medical management of hypertension of hypertension of the study participants in Hawassa City, Ethiopia, 2023 (n=300)

| Variables | Frequency | Present (%) |
| --- | --- | --- |
| Know the normal range of BP measurements |  |  |
| Yes | 261 | 87.0 |
| No | 39 | 13.0 |
| Hypertension can be preventable |  |  |
| Yes | 260 | 86.7 |
| No | 40 | 13.3 |
| Ever heard about the non-medical management of HTN |  |  |
| Yes | 183 | 61.0 |
| No | 117 | 39.0 |
| Know how to reduce the risk behaviours of HTN |  |  |
| Yes | 249 | 83.0 |
| No | 51 | 17.0 |
| Know the importance of maintaining normal body weight to control BP |  |  |
| Yes | 263 | 87.7 |
| No | 37 | 12.3 |
| Know the importance of diet in controlling BP |  |  |
| Yes | 275 | 91.7 |
| No | 25 | 8.3 |
| Know the importance of reducing salt intake in controlling BP |  |  |
| Yes | 298 | 99.3 |
| No | 2 | 0.7 |
| Know it's good to have whole fruits and vegetables to lower BP |  |  |
| Yes | 249 | 83.0 |
| No | 51 | 17.0 |
| Know the effect of drinking excessive alcohol on controlling BP |  |  |
| Yes | 237 | 79.0 |
| No | 63 | 21.0 |
| Know the effect of cigarette smoking on controlling BP |  |  |
| Yes | 196 | 65.3 |
| No | 104 | 34.7 |
| Know the importance of regular physical exercise in controlling BP |  |  |
| Yes | 259 | 86.3 |
| No | 41 | 13.7 |
| Know the effect of chewing Khat on controlling BP |  |  |
| Yes | 177 | 59.0 |
| No | 123 | 41.0 |
| Know the effect of caffeine on controlling blood pressure |  |  |
| Yes | 201 | 67.0 |
| No | 99 | 33.0 |
| Overall knowledge (Computed) |  |  |
| Good | 224 | 74.7 |
| Poor | 76 | 25.3 |

### Knowledge level of respondents towards non-medical management of hypertension

In this study, the overall proportion of participants with a good Knowledge level towards non-medical management of hypertension was found to be 74.7 % (n=224). Regarding specific knowledge about controlling hypertension, 83% (n=249) knew how to reduce the risk behaviours associated with HTN, and 87.7% (n=263) recognized the importance of maintaining a normal body weight to control blood pressure. Moreover, 91.7% (n=275) understood the significance of diet in controlling blood pressure, while 99.3% (n=298) acknowledged the importance of reducing salt intake for BP control. Participants were also knowledgeable about the beneficial effects of certain practices, as 83% (n=249) knew that consuming whole fruits and vegetables could help lower blood pressure. Furthermore, 79% (n=237) were aware of the adverse effects of excessive alcohol consumption on blood pressure, and 65.3% (n=196) understood the impact of smoking cigarettes on blood pressure. Additionally, 86.3% (n=259) were aware of the importance of regular physical exercise in controlling blood pressure, while 59% (n=177) recognized the effects of chewing khat on blood pressure. Lastly, 67.0% (n=201) were knowledgeable about the effects of caffeine on controlling blood pressure. (Table 5).

**Table 5:** Bivariate and multivariate analysis of factors associated with practices of non-medical management of hypertension, Hawassa City, Ethiopia.

| Variables | Practices of NMMs of HTN |  | COR (95% CI) | AOR (95% CI) | P-Value |
| --- | --- | --- | --- | --- | --- |
|  | Good | Poor |  |  |  |
| Permanent resident |  |  |  |  |  |
| Urban (ref) | 142(58.4%) | 101(41.6%) |  |  |  |
| Rural | 45(78.9%) | 12(21.1%) | 2.667(1.343, 5.296) | 1.939 (0.943, 3.987) | 0.072 |

| Level of knowledge |  |  |  |  |  |
| --- | --- | --- | --- | --- | --- |
| Good | 152(67.9%) | 72(32.1%) | 2.473(1.454, 4.206) | 2.368 (1.354, 4.141) | **0.003** |
| Low(ref) | 35(46.1%) | 41(53.9%) |  |  |  |
| Level of Attitude |  |  |  |  |  |
| Favourable | 81(77.1%) | 24(22.9%) | 0.353(0.207, 0.603) | 2.054 (1.180, 3.575) | **0.011** |
| Unfavourable (ref) | 106(54.4%) | 89(45.6%) |  |  |  |
| Family History Of Hypertension |  |  |  |  |  |
| Yes | 54(45.4%) | 65(54.6%) | 1.718 (1.067, 2.766) | 1.764 (1.073, 2.899) | **0.025** |
| No(ref) | 59(32.6%) | 122(67.4%) |  |  |  |

### Health profile-related characteristics

Of the study participants, 39.7 %(n=119) had a family history of HTN and 27.7 % (n=83) of the respondents had co-morbidities. (Figure-1)

**Figure-I:**
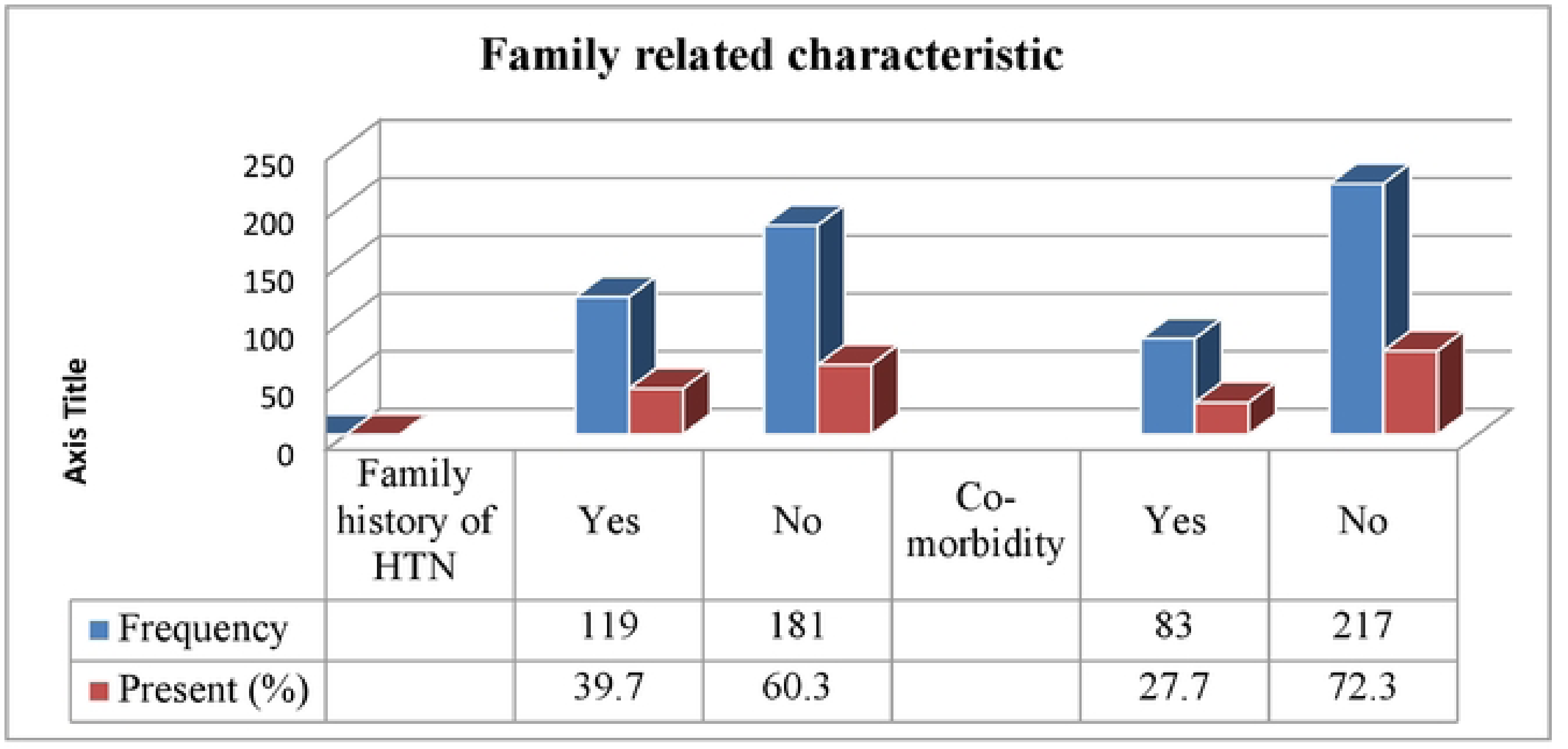
Health profile related characteristic of hypertension of the study participants in Hawassa city, Ethiopia, 2023 (n=300)

### Factors Associated with Practices of non-medical management of hypertension

Logistic regression analysis was conducted to assess factors associated with practices of non-medical management of hypertension. The Variables included in the analysis were rural residence, good knowledge level, favourable attitude towards non-medical management of hypertension and a family history of hypertension. As a result, the level of knowledge, and attitude towards non-medical management of hypertension and having a family history of hypertension were found to be significantly associated with practices of non-medical management of hypertension.

This study revealed that rural residents had higher odds of practising NMM practice of hypertension compared to urban residents (AOR 1.939, 95% CI: 0.943-3.987), P-value=0.072. Respondents with good knowledge level toward practices of non-medical management of hypertension among respondents were approximately two times more likely to practice these strategies compared to those with low knowledge (AOR 2.368, 95% CI: 1.354-4.141), p-value=0.003) Similarly, respondents with a favourable attitude towards non-medical management of hypertension had around two times higher odds of practising these methods compared to those with an unfavourable attitude (AOR=2.054, 95% CI: 1.180-3.575, P-value=0.011). Furthermore, respondents who had a family history of hypertension were approximately two times more likely to practice non-medical management strategies compared to those without a family history of hypertension (AOR=1.764, 95% CI: 1.073-2.899, P-value=0.025). These findings indicate that both knowledge and attitude towards non-medical management of hypertension, as well as having a family history of hypertension, are significant factors influencing the adoption of non-medical management practices among the respondents. (Table-5)

## Discussion

In this study, the prevalence of good practices towards non-medical management of hypertension among the study participants was found to be 62.3%. This indicates that nearly two-thirds of hypertensive patients had good practices towards non-medical management of hypertension. This finding is higher than the study conducted in Addis Ababa, the level of good practice to recommended LSM was found to be 23% (11), in West Aresi zone, Oromia 25.2% (9), in Mizan Tepi 33.3% (12), a study in Durame, southern Ethiopia was 27.3% (13), in central Gondar zone, the overall prevalence of recommended lifestyle modification was 24.2% (14), in Dessie, northeast Ethiopia, adherence to lifestyle modifications was 23.6%(15), at Bishoftu General Hospital, of the respondents had good practices was 38% (16) and in Hiwot Fana Specialized University Hospital in Harar, Eastern Ethiopia was 136 (49.6%) had good practice on lifestyle modification (10).

On the other hand, this finding is similar to the study conducted in Bishoftu General Hospital, among the total respondents, 72.27% have good knowledge, 68.32% have a good attitude and 61.39% have good practice (17), This finding lower than the study conducted in Nekemte, 68.92% of the respondent had a good practice (7). good knowledge about healthy lifestyle (AOR:0.42; 95% CI: 0.24-0,74) (8), Of the total participants, only 25.2% (95% CI: 18.8-32.9) of the patients practised recommended lifestyle modifications, knowledge on hypertension management (AOR = 14.6, 95% CI: 4.6 - 45.9) (9), 87% of respondents measured their blood pressure regularly compared to 38% of the subjects did not measure their blood pressure regularly and 58.3 % did physical exercise compared to 88% of the study participants did not carry out any form of physical activity (18) Patients who had favorable attitude were 2.054 times more likely practice of NMM than who had unfavourable attitude (AOR: 2.052,95% CI: 1.180, 3.575), patients who had a family history of HTN were 1.764 times more likely practice of NMM than who hadn’t a family history of HTN (AOR: 1.764, 95% CI: 1.073, 2.899)

This study revealed that respondents with a good knowledge level towards non-medical management of hypertension were approximately two times more likely to practice these strategies compared to those with poor knowledge in this area. This finding indicates that hypertensive patients who possessed a better understanding of non-medical management elements were more likely to put these practices into action.

This result is comparable with the study conducted in Ghana, Nigeria (19) and in Dares Salaam, where people who had knowledge of HTN, only 63 (29.6%) had knowledge of the risk factors for HTN (20). On the other hand, respondents with a favourable attitude of the respondents toward non-medical management of hypertension were twofold, more likely to practice these strategies compared to those with an unfavourable attitude. This implies that having a favourable attitude toward the non-medical management of hypertension improves practices of non-medical management of hypertension among patients. This finding was similar to studies conducted in Ghana (19), in Durame, southern Ethiopia, in the Central Gondar zone, in Dessie, northeast Ethiopia (15) and at Bishoftu General Hospital, Oromia Region(8). The differences in the results could be attributed to variations in sample size and sampling methods employed in the respective studies.

On the other hand, respondents with a family history of hypertension were approximately two times more likely to practice non-medical management of hypertension when compared to those without a family history of hypertension. This implies that having a family history of hypertension increases the likelihood practices of non-medical management of hypertension among hypertensive patients. However, this finding is not comparable to some previous studies, possibly due to differences in the study design, sample size, study time, nature of participants and environmental factors. This suggests that having a family history of hypertension increases the likelihood of adopting non-medical management practices among hypertensive patients.

However, this finding is not directly comparable to some previous studies, possibly due to differences in study design, sample size, study setting, study time, nature of study participants, and environmental factors.

## Conclusions and recommendation

This study aimed to assess the practice of non-medical management of hypertension and its associated factors among hypertensive patients. The findings revealed that one-third of the study participants exhibited poor practice in non-medical management of hypertension. Factors such as rural residence, good knowledge level, favourable attitude, and having a family history of hypertension showed statistically significant associations with the practice of non-medical management of hypertension.

Based on these results, it is evident that there is a need for an integrated effort from various health disciplines to enhance patients’ knowledge and attitudes towards non-medical management strategies for hypertension. By addressing these factors and providing comprehensive support, healthcare professionals can play a vital role in improving the overall management of hypertension in patients.

## Data Availability

The data is available from main author for a reasonable request.

## Acknowledgement

We would like to pass sincere thanks to the Hawassa City Administration Health Department and Hospitals for their cooperation in providing information that is important to this study.

Finally, we express our thanks to data collectors and study participants for their time, volunteers, and effort.

